# Machine Learning-Enabled EEG Biomarkers Predict Divergent Antidepressant and Placebo Response in a Clinical Trial of Major Depression

**DOI:** 10.1101/2025.05.29.25328167

**Authors:** Qiang Li, Michael Detke, Steve M. Paul, William Z. Potter, Fan Zhang, Alan Breier, Larry Alphs, Owen M. Wolkowitz, Larry Ereshefsky, Gregory G. Grecco, Ken Wang

## Abstract

**Background:** Major depressive disorder (MDD) is a heterogeneous neuropsychiatric disorder with highly variable antidepressant outcomes. In randomized controlled trials (RCTs), low drug-placebo differences and high placebo response rates are persistent challenges. An objective biomarker that can prospectively identify which patients will respond to antidepressant or placebo could greatly enhance both clinical care and clinical trial outcomes.

**Methods:** Baseline scalp EEG data from EMBARC, a multi-site RCT of the SSRI sertraline vs placebo in adult MDD, were analyzed using unsupervised machine learning to identify subtypes and compare these with their corresponding treatment response profiles. Subtypes response to sertraline versus placebo was evaluated by 8-week HAMD-17 outcomes (change from baseline).

**Results:** Of the 215 subjects, three EEG clusters yielded four response phenotypes. (1) Drug–Responders exhibited a large sertraline advantage over placebo (n = 124; *d* = 1.23; *p* < 0.0001). (2) Non–Responders derived no benefit from sertraline (n = 37; *d* = –0.07; *p* = 0.84). (3) Divergent–Responders shared a distinctive connectivity profile clearly separable from phenotypes 1 and 2. Within this group, participants randomized to placebo improved robustly (Placebo–Responders; n = 54; *d* = –1.52; *p* < 0.0001), whereas those receiving sertraline worsened (Adverse Drug–Responders; n = 31; *d* = -0.67; *p* = 0.004). Excluding Placebo–Responders more than tripled the overall drug–placebo effect size (*d* = 0.89 vs 0.28). Cluster membership was highly stable in 10–fold cross–validation (98–99 % consistency) and reproduced across three independent trial sites, underscoring generalizability.

**Conclusions:** Scalp EEG activity analyzed with machine learning identified four biomarker-defined subtypes with strikingly distinct responses to an antidepressant and placebo. These results raise the possibility of using low-cost, noninvasive EEG to guide personalized treatment decisions, avoid ineffective or harmful medications, and improve clinical trial outcomes by identifying drug and high placebo responders in advance of initiating treatment.

## Introduction

Major depressive disorder (MDD) is a common, disabling and potentially fatal neuropsychiatric disorder with a lifetime prevalence rate of 20% in the U.S. [1]. More than 300 million people worldwide are living with depression, making it a leading cause of global disability and death [2]. Approximately 60% of suicides in the U.S., a leading cause of death in several age groups, are due to untreated or inadequately treated depression [3]. Standard treatments for MDD, including antidepressant medications (most commonly selective serotonin reuptake inhibitors, SSRIs, and similar agents), psychotherapy, and neuromodulation, produce highly variable outcomes. The clinical heterogeneity of depression means that any given treatment may only work for a subset of patients. Prescribing an antidepressant is often a “trial and error” process; clinicians empirically choose a therapy and wait weeks to observe a response [4]. This delay (clinical trials of antidepressants typically last 6–12 weeks) not only prolongs suffering but also carries risk, as ongoing depression is associated with impaired functioning and a heightened risk of suicide [5, 6]. An objective, pre-treatment predictor of antidepressant response or non-response could revolutionize care by enabling personalized treatment selection. If one could identify likely non-responders or those prone to adverse effects before starting a medication, alternative strategies (switching drug class, psychotherapy, etc.) could be pursued sooner. Likewise, identifying patients who are likely to respond well to a given antidepressant would instill confidence in continuing a given course of treatment.

Beyond individual patient care, depression’s heterogeneity and the placebo response complicate and confound clinical trials for new treatments. Randomized controlled clinical trials (RCTs) for antidepressants often show that antidepressants outperform placebo by a standardized mean difference of only about 0.3 in symptom reduction [7], which may be statistically significant in large samples but is otherwise clinically quite modest. High placebo response rates (often 30–40% of patients improve on placebo) are one of the most vexing challenges in demonstrating the efficacy of novel antidepressants [8, 9]. Despite decades of exploring methods for reducing the placebo response, such as placebo “run-in” phases, lead-in psychotherapy, enrichment strategies, or more stringent inclusion criteria, no strategy has yielded a reliably effective solution [9–12]. In fact, despite methodological refinements since the 1980s (when SSRI trials became common), placebo responses remain substantial and show no clear downward trend. Identifying placebo responders before randomization could transform clinical trials by improving drug-placebo differences and reducing sample size requirements.

Biomarkers have long been pursued to predict antidepressant treatment response in depression with EEG and advanced neuroimaging techniques showing significant promise [13–17]. While MRI provides insights into deep-brain structures, its high cost and limited accessibility constrain its clinical utility. In contrast, EEG is inexpensive, noninvasive, and widely available, offering a scalable platform for real-time brain monitoring that aligns with the goals of precision psychiatry. Prior studies have linked various EEG features to treatment outcomes in MDD, though early efforts often struggled to differentiate medication response from placebo effects in prospective settings [14, 18–21]. However, MDD is increasingly understood as a disorder of large-scale brain network dysfunction marked by features such as decreased activity in executive control networks and hyperconnectivity in the default mode network [22–26]. These aberrant connectivity patterns suggest that functional relationships between brain regions, rather than isolated activity in specific regions, may better explain treatment response. By applying machine learning to analyze resting-state EEG connectivity, it may be possible to uncover distinct neurophysiological-defined subtypes of depression that correspond to differential outcomes with antidepressant medication and placebo. Indeed, others have used similar data-driven clustering approaches to identify clinically meaningful depression subtypes, demonstrating utility in predicting which patients will respond to specific antidepressants [27–31].

In the present study, we apply an unsupervised machine learning approach to analyze baseline resting-state EEG from the Establishing Moderators and Biosignatures of Antidepressant Response for Clinical Care for Depression (EMBARC), a multi-site RCT of sertraline vs. placebo in MDD [32], to uncover natural subtypes of depression based on functional connectivity. This study extends upon our prior exploratory findings previously presented [33]. We hypothesized that data-driven clustering of EEG functional connectivity features could reveal neurophysiological subtypes of depression with unique treatment response profiles. We then examined whether these EEG-defined subgroups showed differential response to sertraline versus placebo. We identified responder, non-responder, placebo-responder, and adverse-responder subtypes which, if validated prospectively, could personalize treatment and optimize clinical trial outcomes in MDD.

## Methods

### Study Design and Sample

In this study, we conducted an analysis of baseline resting-state scalp EEG data obtained from EMBARC, a multi-site, double-blind, placebo-controlled RCT of sertraline in patients with MDD [32] retrieved from the NIMH Data Archive (NDA) database. This original study adhered to FDA and Helsinki guidelines and was approved by Institutional Review Boards. In brief, EMBARC recruited outpatients aged 18-65 with current unipolar MDD (baseline HAMD-17 ≥ 14), who had no history of failure on an SSRI, and no contraindications to SSRIs. After screening and baseline assessments (including EEG and MRI), participants were randomized to 8 weeks of sertraline or placebo under double-blind conditions. For complete details on participant inclusion and exclusion criteria, please see Trivedi et al., 2016. Our study utilized scalp EEG data from three out of four sites, specifically Columbia University (CU), the University of Michigan (UM), and the University of Texas Southwestern Medical Center (TX). Data of 62 subjects from Massachusetts General Hospital (MG) was not utilized due to its geodesic EEG montage layout, different from other sites using a 10-10 standard layout, posing challenges for sensor space analysis.

### EEG Acquisition and Preprocessing

Eyes-open resting-state scalp EEG data recorded at baseline (prior to treatment initiation) were utilized in the study. All data was preprocessed by automatic algorithms, including bad epoch rejection, bad channel removal, and interpolation. Subjects with fewer than 5 good epochs or more than 20% of bad channels were excluded from the study. Table 1. shows the demographic information of the included subjects. The final sample size for analysis was 215 out of 245 randomized MDD subjects, after EEG QC.

**Table 1.** Demographic and Clinical Characteristics Stratified by EEG Signature Group (ESG)

| Variable | ESG-1 (n = 124) | ESG-2 (n = 37) | ESG-3 (n = 54) | p-value | BH q-value |
| --- | --- | --- | --- | --- | --- |
| Age, mean (SD), years | 40.3 (13.2) † | 32.4 (12.0) | 34.4 (13.3) | 0.0006 | 0.002 |
| Female, n (%) | 79 (63.7 %) | 27 (73.0 %) | 43 (79.6 %) | 0.207 | 0.230 |
| Race, n (%) |  |  |  | 0.153 | 0.298 |
| White | 85 (68.5 %) | 19 (51.4 %) | 39 (72.2 %) |  |  |
| Black | 23 (18.5 %) | 9 (24.3 %) | 7 (13.0 %) |  |  |
| Asian | 5 (4.0 %) | 6 (16.2 %) | 4 (7.4 %) |  |  |
| Other | 11 (8.9 %) | 3 (8.1 %) | 4 (7.4 %) |  |  |
| Ethnicity, Hispanic (%) | 25 (20.2 %) | 7 (18.9 %) | 12 (22.2 %) | 0.92 | 0.985 |
| Clinical Site, n (%) |  |  |  | 0.017 | 0.083 |
| CU | 39 (31.5 %) | 19 (51.4 %) | 21 (38.9 %) |  |  |
| TX | 58 (46.8 %) | 6 (16.2 %) | 18 (33.3 %) |  |  |
| UM | 27 (21.8 %) | 12 (32.4 %) | 15 (27.8 %) |  |  |
| Baseline HAMD, mean (SD) | 18.4 (4.7) | 17.7 (4.1) | 17.9 (4.4) | 0.608 | 0.873 |
| Age of MDD Onset, mean (SD), years | 16.3 (6.2) | 15.4 (5.0) | 16.2 (6.0) | 0.753 | 0.873 |
| Number of MDEs, median (IQR) | 4 (2 – 20) | 3 (2.5 – 6.5) | 3 (2 – 5.5) | 0.270 | 0.450 |
| Duration Current MDE, median (IQR), weeks | 20 (5 – 60) | 19 (5 – 36) | 12 (5 – 46.5) | 0.481 | 0.735 |
| GAD, % Yes | 12.1 % | 16.2 % | 17.0 % | 0.897 | 0.985 |
| Panic Disorder, % Yes | 14.2 % | 19.4 % | 18.5 % | 0.149 | 0.298 |
| Social Phobia, % Yes | 17.7 % | 18.9 % | 18.5 % | 0.875 | 0.985 |
| OCD, % Yes | 3.2 % | 5.4 % | 5.7 % | 0.315 | 0.525 |
| PTSD, % Yes | 19.5 % | 16.2 % | 13.0 % | 0.058 | 0.173 |
| Anorexia Nervosa, % Yes | 2.4 % | 0.0 % | 1.9 % | 0.782 | 0.945 |
| Bulimia Nervosa, % Yes | 0.8 % | 0.0 % | 5.6 % | 0.225 | 0.487 |
| Alcohol, % Yes | 18.5 % | 24.3 % | 16.7 % | 0.540 | 0.811 |
| Cannabis, % Yes | 8.9 % | 10.8 % | 7.4 % | 0.952 | 0.985 |
| Stimulants, % Yes | 4.1 % | 2.7 % | 0.0 % | 0.540 | 0.811 |
| Opioid, % Yes | 0.8 % | 0.0 % | 0.0 % | — | — |
| Cocaine, % Yes | 3.3 % | 5.4 % | 3.7 % | 0.967 | 0.985 |
| Hallucinogen/PCP, % Yes | 0.0 % | 0.0 % | 1.9 % | 0.460 | 0.735 |
| Polysubstance, % Yes | 0.8 % | 0.0 % | 0.0 % | — | — |

† Tukey post-hoc: ESG1 significantly older than ESG2 ( $p=0.0028$ ) and ESG3 ( $p=0.0135$ ). For GAD, values represent current GAD co-morbidity, for all other co-morbidities, values represent self-reported lifetime prevalence of disorder, SD, standard deviation; n, sample size; IQR, interquartile range; HAMD, 17-item Hamilton Depression Rating Scale; MDD, major depressive disorder; MDE, major depressive episode; GAD, generalized anxiety disorder; OCD, obsessive-compulsive disorder; PTSD, post-traumatic stress disorder; PCP, phencyclidine (hallucinogen); CU, Columbia University Medical Center; TX, University of Texas Southwestern Medical Center; UM, University of Michigan.

### EEG Connectivity Features

To characterize each patient’s baseline brain network function, sensor-space functional connectivity, which measures the relationship of time course signals between the scalp electrodes without mapping the signals to specific brain regions, was derived for each subject. Three types of pair-wise functional connectivity metrics were used: Covariance with Generalized Eigenvalue Decomposition, Coherence, and Power Envelope Correlation.

### Machine Learning Clustering

Following derivations of the connectivity matrix, clustering algorithms, the K-means clustering algorithm, a form of unsupervised machine learning, were employed to divide the subjects into three clusters based on the patterns of the functional connectivity matrix, referred to as EEG signatures. Since each cluster represents a group of subjects with the same EEG signatures, in the following text, “cluster” will be referred to as “EEG signature group” (ESG-1, ESG-2, and ESG-3).

Based on the fact that different features capture different characteristics of the electrophysiological processes reflective of EEG activity, we put multiple machine-learning pipelines into an Ensemble machine-learning model, as illustrated in **Figure 1**. Multiple machine-learning models were used in parallel, and their outcomes were then combined to generate the final outcome. The strength of this approach is that each model may be sensitive to certain aspects of the original data and may also be sensitive to a certain type of noise. Combining multiple models serves to capture a more representative picture of the data and to suppress noise.

**Figure 1.**
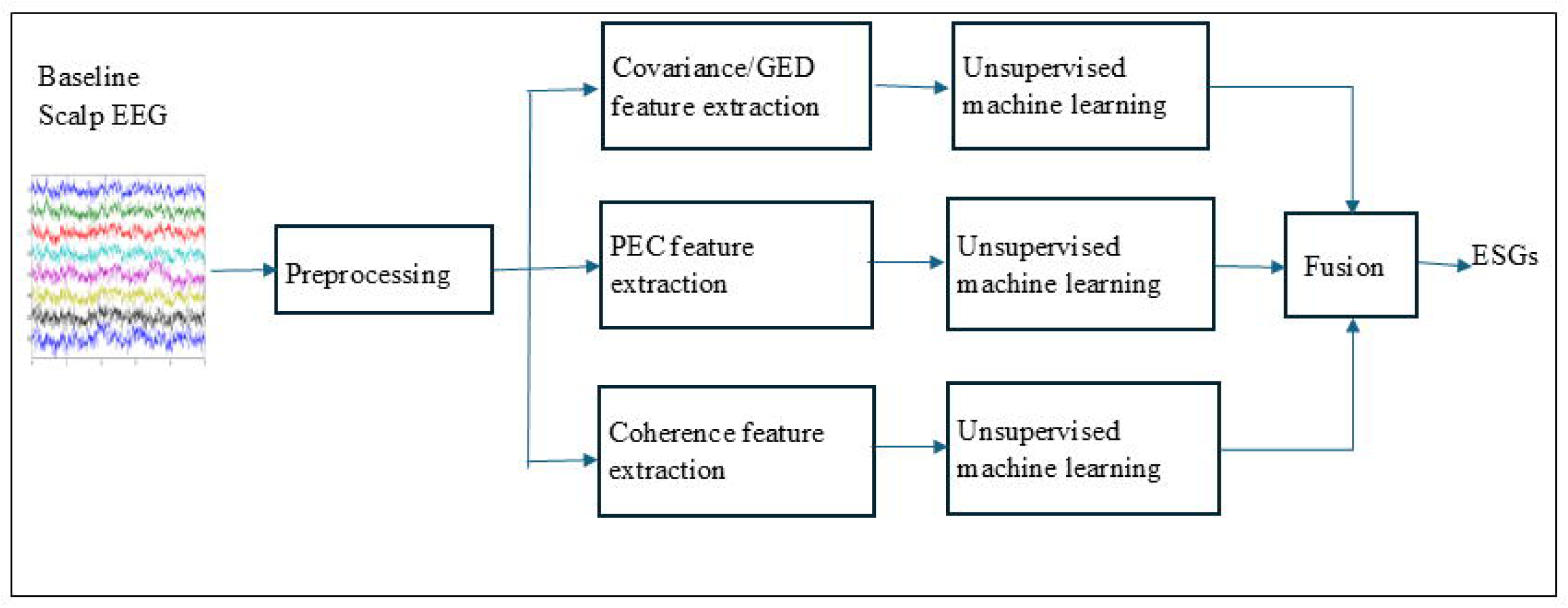
Overview of the ensemble machine learning pipeline used to generate EEG signature groups (ESGs). Baseline resting-state scalp EEG undergoes preprocessing before three distinct feature extraction methods are applied: covariance/generalized eigen decomposition (GED), phase envelope correlation (PEC), and coherence analysis. Each resulting feature set is independently analyzed using unsupervised machine learning to identify EEG-based subgroups. The final ESG assignment is determined through a consensus voting mechanism that integrates the outputs of all three unsupervised models.

Each pipeline assigned subjects to one of three ESGs based on its own clustering results. Cohen’s *d* and p values were calculated for each ESG with regard to any differences between sertraline and placebo. We call the ESG with the greatest *d* in favor of sertraline ESG-1, the least *d* (including negative d) as ESG-3, and the remaining as ESG-2. Then a voting process was carried out. When 2 or more out of 3 pipelines voted a subject to a given ESG, that ESG became the final ESG of that subject. When a subject received 3 different votes, it was assigned to ESG 2.

### Statistics

Differences in average reductions in HAMD-17 total scores, from baseline to the end of the first phase of the trial (Week 8), between the active treatment (sertraline) and placebo arms, were used to assess each ESG, quantified by Cohen’s *d* effect size and the corresponding *p*-value using the two-sample *t*-test. Results at *p* < 0.05 were considered statistically significant. When the Week 8 score was missing, the Week 6 value served as the last observation carried forward (LOCF); participants lacking both Week 8 and Week 6 ratings were omitted from the Cohen’s *d* and *p*-value calculations (30 excluded; 8 imputed). The excluded subjects were still included in the EEG-based subtyping process since their EEG data were independent of the clinical endpoint measurement. The same effect-size and significance tests were applied to each individual HAMD item to explore symptom-level patterns. Baseline demographic and clinical variables were compared across EEG–defined subtypes (ESG 1–3) to characterize potential confounds. Data were first cleaned by coercing non–numeric entries to NaN and recoding the SCID placeholders 888/999 as missing. Continuous variables were examined with the Shapiro–Wilk test (normality) and Levene’s test (homogeneity of variance). If both assumptions held, a one–way ANOVA was used; otherwise the non–parametric Kruskal–Wallis test was applied. When an ANOVA was significant, pair–wise differences were explored with Tukey’s honestly significant difference (HSD) test, which controls the family–wise error rate for those three contrasts. Categorical variables were analyzed with Pearson’s χ² test; when any expected cell count was < 5, categories were collapsed or the exact test was omitted. For SCID lifetime comorbidities, all three response levels (1 = absent, 2 = sub–threshold, 3 = threshold) were entered into a 3 × 3 χ² table, and the percentage of threshold–positive (code 3) cases was reported. Twenty omnibus baseline/comorbidity *p*–values were subjected to a Benjamini–Hochberg false–discovery–rate (FDR) correction (q = 0.05). All processing and statistical analyses were performed in Python 3.9 using MNE 1.7.1, SciPy 1.11.4, scikit–learn 1.3.2, statsmodels 0.14.0, pandas 2.1.4, NumPy 1.26.4.

## Results

### EEG Signature Group Subtypes

Among the 215 subjects included in the EEG analysis after outlier removal (107 on sertraline, 108 on placebo), the unsupervised learning approach revealed three robust EEG clusters (ESG-1, ESG-2, ESG-3), which corresponded to strikingly different clinical outcomes (**Figure 2**). We further categorized these into four intuitive responder subtypes as described below:

**Figure 2.**
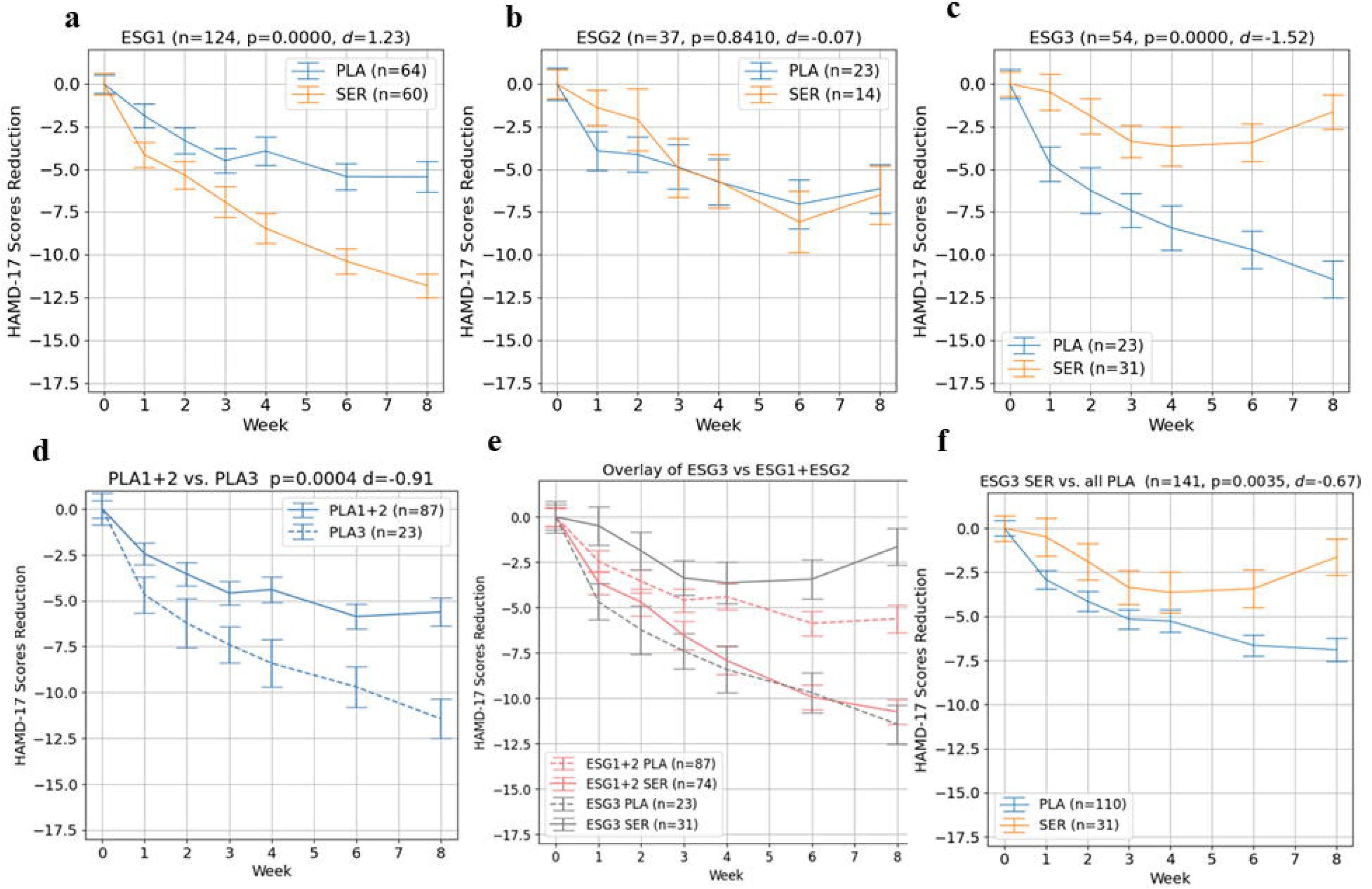
Differential treatment responses across EEG signature groups (ESGs) measured by changes in HAMD-17 scores over 8 weeks. (a) ESG-1 Drug-Responders (n=124): Patients receiving sertraline (SER) exhibited significantly greater symptom reduction than those receiving placebo (PLA) (p<0.00001, Cohen’s d=1.23). (b) ESG-2 Non-Responders (n=37): No significant difference in symptom reduction between SER and PLA groups (p=0.8410, d=-0.07). (c) ESG-3 Divergent-Responders (n=54): Patients in the PLA group demonstrated significantly greater symptom improvement compared to those receiving SER (p<0.00001, d=-1.52). (d) Comparison of PLA-treated participants in ESG-3 (n=31) vs. PLA-treated participants from ESG-1 and ESG-2 (n=93) indicates enhanced placebo response in ESG-3 (*p*=0.0004, *d*=-0.91). (e) Overlay of mean symptom trajectories for ESG-3 and the combined ESG-1 & ESG-2 subtypes, highlighting distinct patterns of response to placebo. (f) ESG-3 SER group (n=23) compared to all PLA-treated participants (n=110), showing inferior outcomes for sertraline in ESG-3 (*p*=0.0035, *d*=-0.67). Error bars represent standard error of the mean (SEM).

#### Subtype 1-Strong SSRI Responders (ESG-1)

This was the largest cluster, comprising 124 patients (58% of the sample). Patients in Subtype 1 showed a marked benefit from sertraline compared to placebo. Their average HAMD-17 scores over 8 weeks improved much more on sertraline (mean reduction ≈ –11 points) than on placebo (mean reduction ≈ –6 points). The drug-placebo difference for Subtype 1 was highly significant (Cohen’s *d* = 1.23 in favor of sertraline, *p* < 0.0001; **Figure 2a**). This finding is noteworthy because typical MDD trial populations rarely exhibit such a large effect size; most trials assume an expected *d* ∼0.3 for drug vs placebo [7]. Here, using pretreatment EEG, we identified a sizable subset of patients with an effect size roughly four times greater than average.

#### Subtype 2 - Non-Responders (ESG-2)

This cluster included 37 patients (17% of the sample) and was characterized by minimal improvement in both treatment arms. Patients in Subtype 2 had relatively flat depression trajectories, whether they received sertraline or placebo (**Figure 2b**). By week 8, the mean HAMD change was small with no significant difference between sertraline and placebo (*d* = –0.07, *p* = 0.841; **Figure 2b**). Clinically, this cluster aligns with the concept of patients who might need alternative treatments (e.g. different mechanisms, combination treatments, etc.), as neither medication nor placebo-associated factors (expectancy, trial participation effects) produced much improvement. Identifying such patients prospectively could help avoid ineffective first-line treatment trials.

The third pretreatment EEG cluster (ESG-3: Divergent Responders made up of Placebo Responders and Adverse Drug Responders) contains 54 patients (25 % of the total) who share a distinctive pretreatment EEG pattern. Although electrophysiologically homogeneous, the cluster “diverges” once treatment is assigned, splitting into two opposite clinical trajectories, hence the label Divergent Responders. These contrasting outcomes create two subtypes:

#### Subtype 3 - Placebo Responders (part of ESG-3)

ESG-3 is comprised of 54 patients (25% of the total) which can be subdivided by treatment: 23 patients in this cluster received placebo (Subtype 3) and 31 received sertraline (Subtype 4, described below). Subtype 3 patients (Placebo Responders) exhibited a remarkable improvement on placebo, comparable to the best drug responders in Subtype 1. Their average HAMD scores dropped by ∼11-12 points on placebo, indicating a near remission-level response; whereas, had they been given sertraline, their improvement would likely have been significantly smaller (as evidenced by Subtype 4 below). Within ESG-3, the difference between the placebo arm and the drug arm was striking as placebo outperformed sertraline with a very large effect size (*d* = -1.52, *p* < 0.00001; **Figure 2c**). Moreover, when comparing Subtype 3’s placebo response to all other placebo patients (i.e. placebo-treated patients not in ESG-3), Subtype 3 clearly did better (Cohen’s *d* = -0.91 vs the rest of placebo group, *p* = 0.0004; **Figure 2d**). Therefore, the pretreatment EEG signature defining ESG-3 appears to identify individuals who are exceptionally sensitive to placebo effects so much so that they improve as if on active medication. This subset can be considered “Placebo Responders” in the true sense, as their improvement appears to derive largely from placebo and context. Clinically, these could be patients without proper MDD, but who were perhaps undergoing temporary psychosocial stressors at their worst time, or possibly “professional patients” who also do not have MDD but may enroll in clinical trials for attention or minimal financial compensation.

#### Subtype 4 - Adverse Drug Responders (other half of ESG-3)

The remaining 31 patients in ESG-3 received sertraline, and paradoxically they had the worst outcomes of any group. This Subtype 4 can be described as “Adverse Drug Responders” to the SSRI, since they improved less on sertraline than they likely would have on placebo. On average, Subtype 4 patients showed a negligible or poor response to sertraline (mean HAMD reduction ∼–3), and some even had symptom worsening. Comparing Subtype 4’s sertraline outcomes to the placebo outcomes of Subtype 3, we observed an effect size of *d* = –1.52 (**Figure 2c**) within the same EEG cluster. When comparing Subtype 4’s sertraline response to the placebo response of all other patients outside ESG-3, sertraline was significantly worse (*d* = –0.67, *p* = 0.0035; **Figure 2f**). These patients not only failed to benefit from the drug, but likely would have done better had they not received it.

### Baseline Characteristics Across EEG Subtypes

Baseline demographic and clinical characteristics were largely indistinguishable across the three EEG–defined subtypes (**Table 1**). After correcting 20 omnibus tests with the Benjamini–Hochberg procedure (*q* = 0.05), only age remained significant (ESG-2 and ESG-3 were on average ≈ 6-8 years younger than ESG-1). All other variables, including sex distribution, race/ethnicity, clinical–site mix, depression severity and chronicity, and 14 SCID comorbidities, showed no subtype differences (*q* > 0.05). Thus, conventional clinical metrics would not have clearly separated these groups; the subtype distinctions emerge only when pretreatment resting–state EEG connectivity is considered, underscoring its added value for identifying biologically meaningful patient sub–populations.

### Impact of Subtype Stratification on Trial Outcomes

The effect of removing high placebo responder (ESG-3) subjects on an SER-PLA comparison of all remaining members from ESG-1 and ESG-2 was evaluated. In the EMBARC trial as a whole (all-comers analysis without stratification), sertraline did not separate significantly from placebo on the primary outcome at 8 weeks (a common outcome in antidepressant trials due to placebo effects). In our complete EEG subset (N=215), the overall drug-placebo effect size was *d* = 0.275 with *p* = 0.065 (**Figure 3a**). However, using our biomarker enrichment strategy, it is possible to remove ESG-3 (i.e. a “screen-failed” cluster of likely Placebo-Responders and Adverse Drug Responders based on pretreatment EEG; **Figure 2c**) which equates to approximately 25% of the total subjects. This results in a screen-success group of 75% of the candidates with sertraline now significantly outperforming placebo in the enriched sample (Cohen’s *d* = 0.89, *p* < 0.0001; **Figure 3b**).

**Figure 3.**
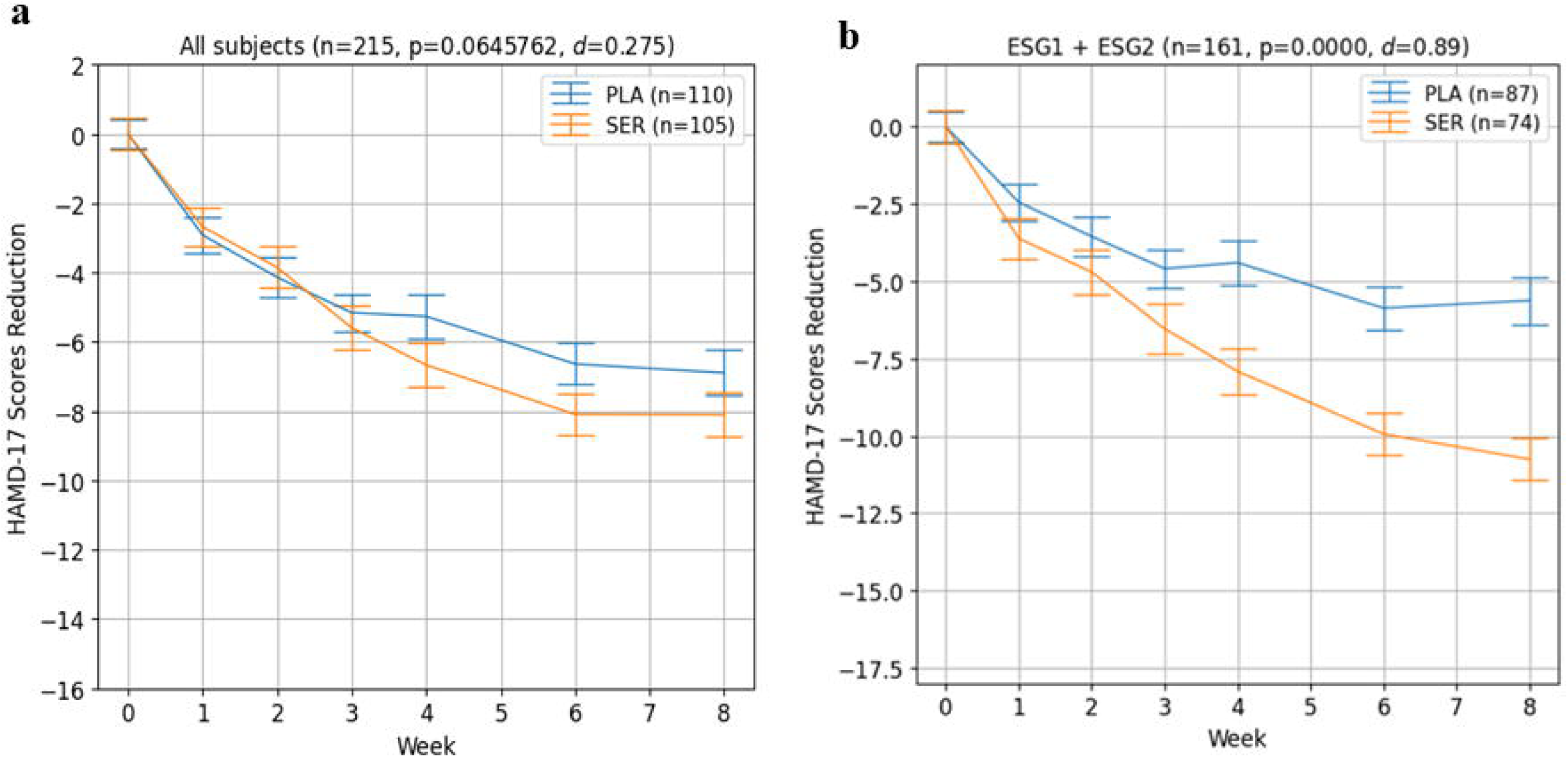
Effect of EEG-based subtyping on treatment signal detection. (a) In the full sample (n=215), there was no statistically significant difference in HAMD-17 score reduction between the sertraline (SER, n=105) and placebo (PLA, n=110) groups (*p*=0.065, *d*=0.275). (b) After excluding ESG-3 and analyzing only ESG-1 and ESG-2 (n=161), a significant treatment effect favoring sertraline emerged (*p*=0.0000006, *d*=0.890), demonstrating the utility of EEG-based subtyping to enrich the sample and uncover a robust drug-placebo separation. Error bars represent standard error of the mean (SEM).

Focusing on just patients who were on sertraline as in an open-label study, a comparison of those who fell into ESG-1 and those in combined ESG-2 and ESG-3 revealed that the difference between high and low sertraline responders (**Figure 4**; *p* < 0.00001, *d* = 1.43) was of an even larger effect size as was observed between sertraline and placebo in the ESG-1 comparison (**Figure 2a**).

**Figure 4.**
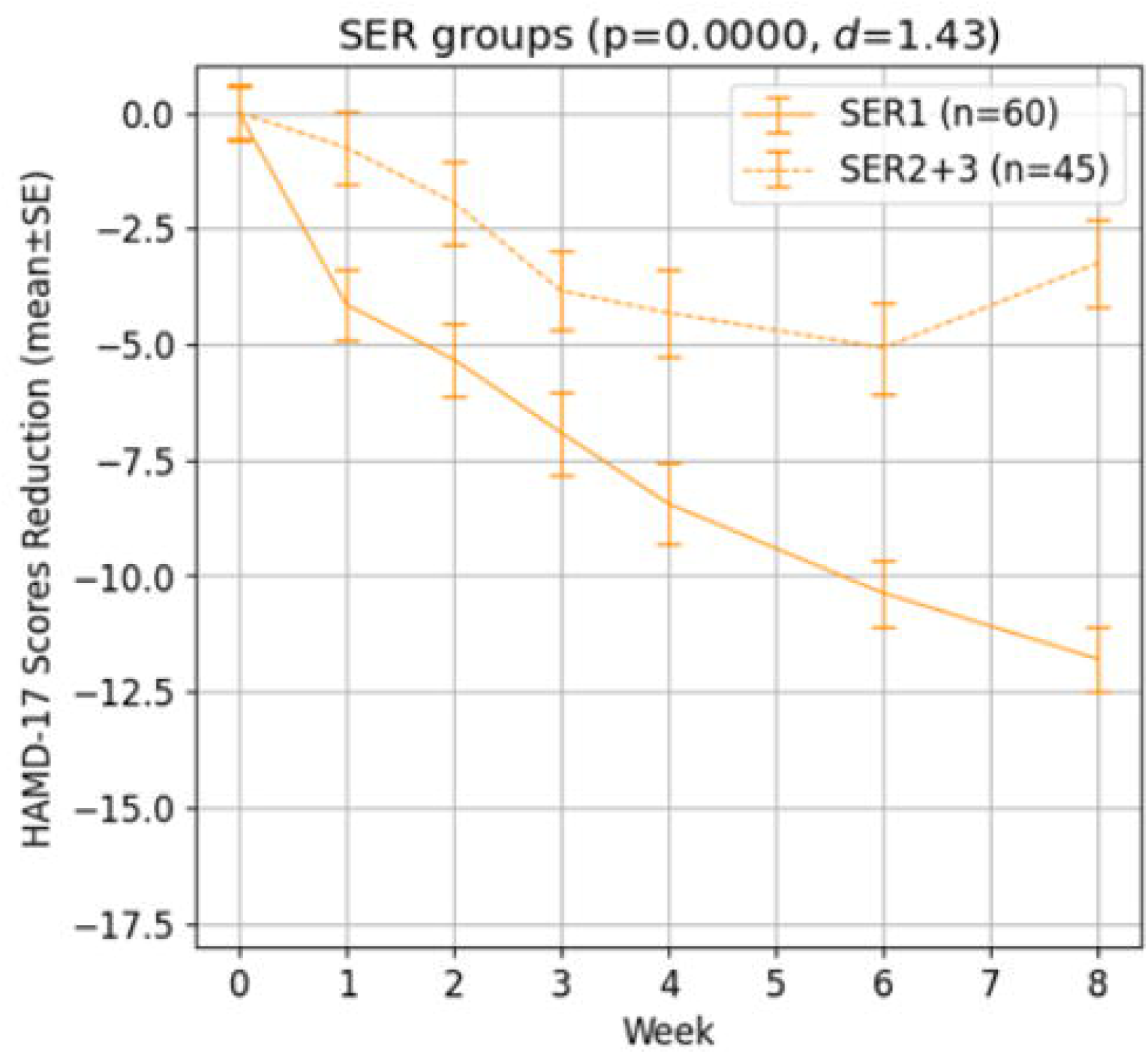
Sertraline treatment response stratified by biomarker-defined subgroups. EEG Signature Group 1 (ESG-1), designated as biomarker-positive, demonstrated significantly greater symptom reduction over 8 weeks compared to the biomarker-negative group (combined ESG-2 and ESG-3; *p*<0.00001, *d*=1.43). Solid line represents the sertraline-treated biomarker-positive group (n=60); dashed line represents the sertraline-treated biomarker-negative group (n=45). Error bars represent standard error of the mean (SEM).

### Sensor-Space PEC Connectivity Heatmaps

To better visualize the differences for each ESG we displayed the average connectivity strength matrix in the form of a heatmap. Although it is difficult to provide a precise neurophysiological and anatomical correlation to each heatmap, such displays may provide insights for further study.

Visualization of the connectivity differences among ESGs is shown as distinct functional connectivity strength heatmaps in **Figure 5**. The placebo responder and adverse drug responder group of ESG-3, had a very distinct sensor-space connectivity strength heatmap, displaying higher contrast between the high connectivity levels in the frontal cortical areas, and the low connectivity levels in the parietal and posterior areas, while ESG-3 displayed lower contrast between the connectivity levels of the frontal areas and the parietal and posterior areas.

**Figure 5.**
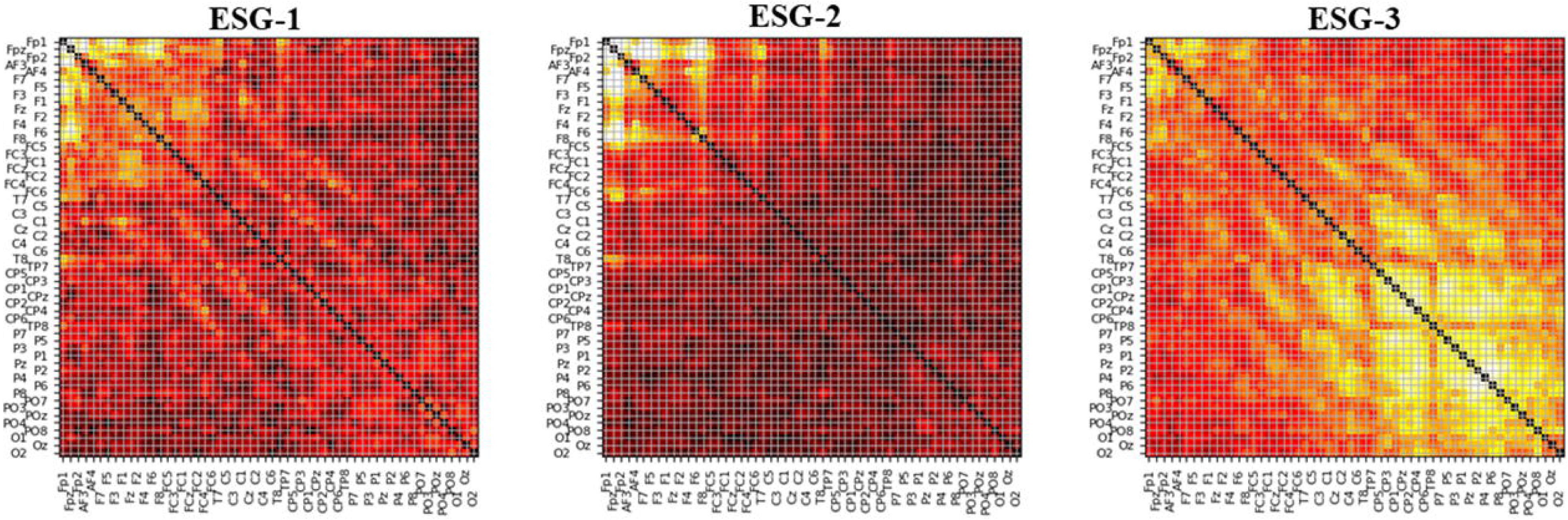
Sensor-space phase envelope correlation (PEC) connectivity heatmaps for each EEG signature group (ESG). Heatmaps were generated by averaging PEC connectivity matrices across all subjects within each subtype (ESG 1-Drug Responders, ESG-2-Non Responders, ESG-3-Divergent Responders). Each matrix is symmetric, with both axes representing EEG channel labels ordered from frontal (top-left) to posterior (bottom-right) regions. Warmer colors (yellow/white) indicate stronger connectivity; cooler colors (dark red/black) indicate weaker connectivity. All heatmaps are normalized to the same color scale to allow direct visual comparison across subtypes.

### Predictive Utility of EEG-Derived Subtypes for Clinical Response

Although not the primary purpose of this investigation, the utility of a predictive test lies in its ability to guide treatment for individual patients, so we next evaluated how well the EEG-derived biomarker could stratify patients based on treatment response which is summarized in **Table 2**. When Subtype 1 was designated as biomarker-positive (**Figure 4**, solid line) and Subtypes 2 and 3 as biomarker-negative (**Figure 4**, broken line), this grouping correctly identified remitters (defined as a final HAMD-17 score ≤ 7) with a sensitivity of 84.3%, specificity of 75.8%, PPV of 75.4%, and NPV of 84.6%. For identifying treatment responders (≥ 50% reduction in HAMD-17 score), the sensitivity and specificity were 84.2% and 79.6%, respectively, with PPV = 82.8% and NPV = 81.3%. These results suggest the EEG-based subtyping approach offers clinically meaningful predictive value using a single predefined threshold.

**Table 2.** Classification Metrics for EEG-Derived Biomarker in Predicting Remission and Response

| **Classification Task** | **Sensitivity (%)** | **Specificity (%)** | **PPV (%)** | **NPV (%)** |
| --- | --- | --- | --- | --- |
| **Remission** | 84.3 | 75.8 | 75.4 | 84.6 |
| **Response** | 84.2 | 79.6 | 82.8 | 81.3 |

### Validation of EEG-Based Subtypes

#### Internal Cross-Validation

K-fold cross-validation was performed to demonstrate that each patient’s cluster assignment remained highly stable. For this, the dataset was randomly partitioned into K approximately equal-sized partitions; K-1 of those partitions were used to train the machine learning model with 1 partition utilized as the testing target. All K combinations were exercised, and the ESG membership of each subject was recorded. This introduced approximately 1/K perturbations to the training data. A 10-fold validation was run 1000 times to evaluate the consistency of subject assignment to their respective ESGs. Results exhibited 98.3% ESG membership consistency for all three ESGs and 99.2% for the placebo responder ESG-3. Such stability is reassuring for potential generalization as the EEG pattern that defines any responder subtype appears to be a clear, reproducible signal that does not depend on any one subset of patients.

#### Cross-Site Validation

EMBARC’s multi-site design allowed us to investigate whether our machine learning model trained with one trial site can produce similar results with data from other trial sites. Cross-site validation is done by training the machine learning model with the data from a single site and testing the model using data from the other two sites, mimicking the scenario where a trained and frozen machine learning model is used in a real-world application with new data. We took advantage of the fact that the EMBARC study was conducted by 4 independent and unaffiliated clinical trial sites (3 sites were included for reasons described earlier) following the same protocol but using different staff and equipment. Cross-site validation provides excellent face validity, as one might expect there would be differences across clinical trial sites with different patient populations, clinicians, local customs, and even weather. In total, 3 models were built with data from CU, TX, and UM, respectively. A representative illustration of this cross-site validation is illustrated in **Figure 6**. For each model, the HAMD-17 score curves for ESG-3 (Placebo and Adverse Responders) are depicted showing compelling consistency across individual, geographically distinct trial sites (**Figure 6**). For all cases, a super placebo responder group was identified as follows: CU model: n = 30 (out of 136 testing subjects, 22%), *d* = -1.31, *p* = 0.0042; TX model: n = 26 (out of 133 subjects, 20%), *d* = -2.5, *p* = 0.000012; and UM model: n = 25 (out of 161 subjects, 16%), *d* = -1.73, *p* = 0.00051. It is noteworthy that the data used for building any single site model was much smaller than the entire trial (e.g. the UM model had only 54 subjects), and yet the models were still able to identify the super placebo responder group, demonstrating the stability of our ensemble model approach.

**Figure 6.**
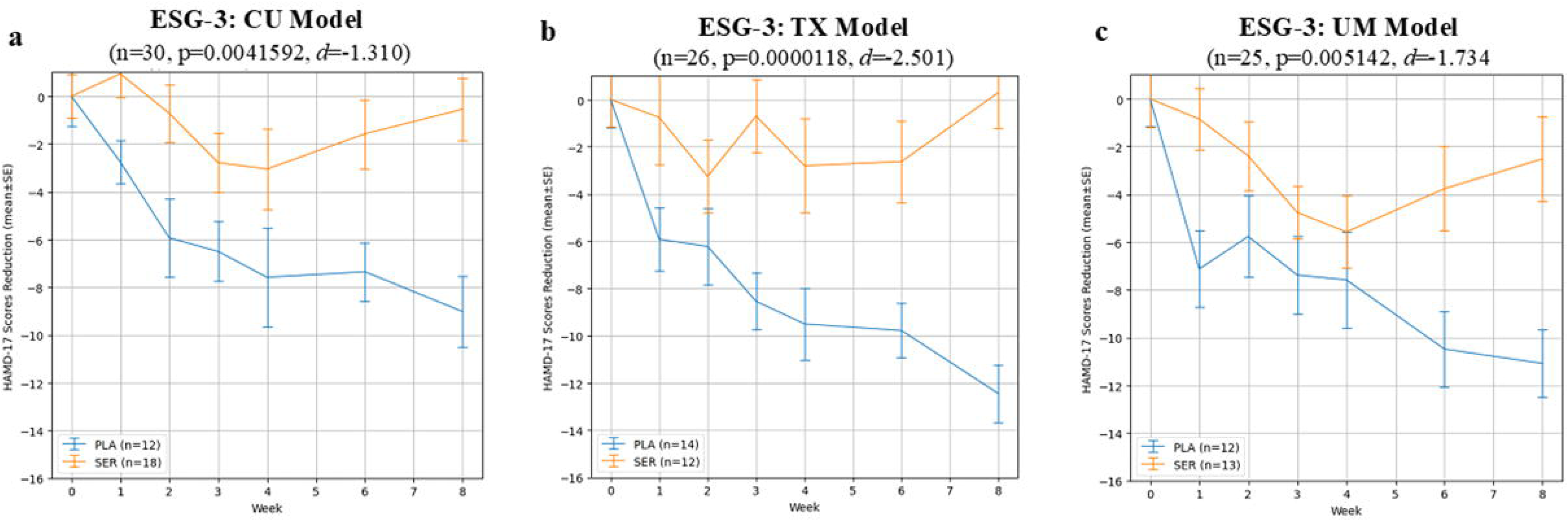
Cross-site validation of EEG-derived signature groups using leave-one-site-out machine learning models. Models were trained on data from one site and tested on held-out subjects from the other two sites. While all EEG signature groups (ESGs) were cross-validated, only ESG-3 is shown here for brevity, as it demonstrated the most clinically striking pattern. Within the ESG-3 Divergent-Responders, the superior placebo response was consistently identified across sites. (a) Model trained on data from Columbia University/Stony Brook (CU), tested on subjects from the University of Texas Southwestern Medical Center (TX) and University of Michigan (UM) (n=30; *p*=0.0042, *d*=1.310). (b) TX-trained model tested on CU and UM subjects (n=26; *p*=0.0001, *d*=2.501). (c) UM-trained model tested on CU and TX subjects (n=25; *p*=0.0051, *d*=1.734). In all panels, placebo-treated participants (blue) showed greater HAMD-17 symptom reduction than those treated with sertraline (orange), consistent with ESG-3’s profile. Error bars represent standard error of the mean (SEM).

## Discussion

In this study, we demonstrate that baseline resting-state EEG biomarkers enabled by machine learning can prospectively identify subtypes of patients with a diagnosis of depression with distinct and clinically meaningful treatment responses in a randomized controlled trial of sertraline versus placebo. Using a data-driven ensemble machine learning approach, we uncovered patterns of EEG functional connectivity that clustered patients into four response profiles: those who responded robustly to sertraline but not placebo (Drug Responders), those who responded to neither (Non-Responders), those who improved substantially on placebo but not drug (Placebo Responders), and those who worsened on sertraline relative to placebo (Adverse Drug Responders). These subtypes were not distinguishable using standard clinical or demographic variables and were validated across multiple independent sites.

Importantly, this stratification was achieved prospectively, using only pre-treatment EEG data, enabling accurate prediction of both response and remission outcomes. Our findings represent an important step toward precision psychiatry for MDD and illustrate that heterogeneity in treatment outcomes is not merely random but is predictable and rooted in measurable neurophysiology.

It is useful to contextualize these results with theoretical response subtypes that clinicians have long suspected. Conceptually, there are theoretically four potential drug or placebo responder subgroups in RCTs of MDD patients: (1) True Drug Responders (2) Non-Responders (3) Placebo-Responders and (4) Adverse-Drug Responders identified as Subtype 4. While each of these patient subtypes has been observed anecdotally or in post-hoc analyses of RCTs, they have never before been identifiable in advance of treatment using an objective biomarker. Given that most clinical trials of antidepressants are powered on the assumption of an effect size of approximately 0.30 [7], to identify populations that showed effect sizes of 1.0 or greater between drug and placebo using EEG connectivity measures would constitute a major advance in clinical trial design. From a clinical trial design perspective, our results are potentially paradigm-shifting. We showed that by excluding the ∼25% of patients who were likely Placebo Responders or Adverse Drug Responders (based on pretreatment EEG), the measured effect size of the drug tripled. This implies that if one were to use such an EEG biomarker as an inclusion/exclusion criterion, trials could achieve the same power with a fraction of the sample size. For example, an effect size of ∼0.9 could mean needing only tens of patients per arm rather than hundreds to confirm efficacy. This could dramatically reduce the cost and duration of antidepressant trials and also reduce participants’ exposure to potentially ineffective experimental drugs. It is worth noting that current approaches to mitigate placebo response (like lead-in periods that drop early improvers) are imperfect and often drop both placebo-responsive and genuinely improving patients [9–12]. In contrast, a biomarker that prospectively identifies placebo-prone individuals would be far more targeted. Our findings echo the call for biomarker-enriched designs in psychiatry, much like oncology has done with genetic markers, to find efficacy signals that are otherwise obscured by heterogeneity [34–37]. A generalizable placebo-response predictor could be invaluable not only in depression trials but in any clinical trial where placebo effects are significant.

Crucially, the EEG biomarker identified was predictive *without* incorporating any overt clinical data. The algorithm did not “know” which patients improved or not; it clustered based purely on baseline brain patterns, yet those patterns turned out to map onto outcome differences. This underscores that there are latent neurobiological signatures in patients diagnosed with depression that carry prognostic information. The topographic EEG connectivity differences between subtypes (visualized in heat maps) hint at underlying brain network disparities. Although we cannot explain the neurophysiological basis of each cluster’s EEG signature, the existence of these signatures is itself instructive. It suggests that certain baseline brain network configurations predispose a patient to respond to the environmental/contextual aspects of treatment (placebo) versus the pharmacological action of an SSRI. For instance, elevated connectivity within the default mode network (DMN) is associated with depressive rumination and poorer antidepressant response, while effective connectivity between DMN and salience networks appears to facilitate symptom improvement across both pharmacological and placebo interventions [38, 39]. Additionally, differences in frontal versus posterior EEG connectivity patterns may further differentiate SSRI responders from placebo responders, reflecting distinct neural mechanisms recruited by medication versus contextual therapeutic effects [40].

It is worthwhile to contrast our EEG-based biomarker findings with fMRI, which has long been considered the gold standard neuroimaging modality in psychiatry and neuroscience. One might assume that fMRI, with its richer spatial detail, or fMRI-based EEG source imaging, which estimates cortical generators of EEG signals, would enhance prediction. However, our preliminary evidence indicates EEG sensor-space analyses outperform MRI in EMBARC (data not shown). This could be due to several reasons. EEG captures millisecond-level neural circuitry interactions; the dynamics of oscillatory connectivity might reflect functional states (like arousal, attention, or network flexibility) that are directly relevant to treatment responsivity whereas fMRI (which measures slower blood-flow changes) might miss these transient but crucial network patterns [34, 41–44]. Additionally, EEG is a more direct measure of brain electrical activity, and perhaps the particular predictive information (for example, a certain pattern of frontal alpha synchrony) is simply not discernible via hemodynamic signals [41, 42]. Our findings align with other work suggesting EEG can be a powerful clinical predictor: for instance, recent meta-analyses have reported EEG-based models achieving ∼80–85% accuracy in predicting antidepressant outcomes [16]. EEG may offer a more accessible and cost-effective alternative to MRI for treatment prediction, with the added advantage of being widely available in routine clinical settings relative for fMRI.

The present EEG ensemble model substantially extends prior EEG biomarker work from the EMBARC study and related analyses, achieving higher predictive accuracy and clearer subtype delineation. While earlier EEG approaches, such as Rolle et al. (2020), identified connectivity patterns that moderated placebo versus sertraline response, they did not provide individually precise classification or robust generalization across diverse sites [40]. Similarly, Zhang et al. (2021) identified two EEG-based subtypes with distinct antidepressant responsiveness, but did not address the issue of placebo responders [45]. Other recent EEG prediction models have also demonstrated only moderate performance (accuracy ∼64%) and modest effect sizes [46, 47]. In contrast, our ensemble method identified four clinically actionable subtypes, including distinct placebo-responder and SSRI super-responder groups, and achieved notably higher predictive accuracy (∼84% sensitivity and specificity) and very large effect sizes (Cohen’s *d*=1.24–1.56). These improvements in accuracy, granularity, and generalizability represent meaningful progress toward individualized and clinically applicable EEG-based treatment stratification for depression.

There are several limitations to our study, however. First, our findings were derived from a single clinical trial dataset of sertraline, albeit one conducted across multiple distinct trial sites with robust validation at each of these sites. While internal and cross-site validation support the initial generalizability of our EEG-defined subtypes, future studies will be required to prospectively replicate these findings in independent datasets, including trials involving other antidepressants, psychotherapies, or placebo-controlled designs, to assess generalizability. Second, the clustering approach required methodological choices such as feature selection and the number of clusters. Although these were guided by objective criteria and ensemble modeling, alternative methods could yield slightly different subgroup boundaries.

Third, our focus on EEG functional connectivity excluded other potentially informative EEG features, such as spectral power or event-related potentials. Future studies could explore whether integrating such features, or adding genetic, behavioral, or other biomarkers, could further refine subtype definitions and predictive performance. Finally, while we observed strong associations between EEG subtypes and treatment outcomes, the neurobiological mechanisms underlying these effects remain unclear. For example, it is unknown why the EEG pattern associated with placebo response emerges in certain individuals. Further studies using additional psychophysiological, cognitive, or molecular assessments may help illuminate the processes that distinguish placebo responders from medication responders.

In summary, this study demonstrates that machine-learning analysis of resting-state EEG can identify biologically distinct subtypes of patients with MDD with divergent responses to antidepressant and placebo treatment. These EEG-derived biomarkers, if validated in independent cohorts, could enable more personalized treatment selection and improve the efficiency of clinical trials by prospectively excluding likely placebo responders. The findings highlight that heterogeneity in treatment outcomes is not random but reflects underlying neurophysiological patterns measurable before treatment begins. Incorporating such biomarkers into clinical practice could represent a major advance toward precision psychiatry for MDD.

## Data Availability

The data analyzed in the present study is publicly available at the NIMH Data Archive. The data produced from current study are not publicly available due to ongoing commercial development. A de-identified limited subset of data may be made available upon reasonable request and subject to a data use agreement.

https://nda.nih.gov/edit_collection.html?id=2199

## References

1. Hasin, D.S., et al., Epidemiology of Adult DSM-5 Major Depressive Disorder and Its Specifiers in the United States. JAMA Psychiatry, 2018. 75(4): p. 336–346.

2. Liu, Q., et al., Changes in the global burden of depression from 1990 to 2017: Findings from the Global Burden of Disease study. Journal of Psychiatric Research, 2020. 126: p. 134–140.

3. Rihmer, Z., et al., Decreasing tendency of seasonality in suicide may indicate lowering rate of depressive suicides in the population. Psychiatry Research, 1998. 81(2): p. 233–240.

4. Leuchter, A.F., et al., A new paradigm for the prediction of antidepressant treatment response. Dialogues Clin Neurosci, 2009. 11(4): p. 435–46.

5. Semkovska, M., et al., Cognitive function following a major depressive episode: a systematic review and meta-analysis. Lancet Psychiatry, 2019. 6(10): p. 851–861.

6. Hawton, K., et al., Risk factors for suicide in individuals with depression: A systematic review. Journal of Affective Disorders, 2013. 147(1): p. 17–28.

7. Curkovic, M., A. Kosec, and A. Savic, Re-evaluation of Significance and the Implications of Placebo Effect in Antidepressant Therapy. Frontiers in Psychiatry, 2019. Volume 10 - 2019.

8. Sonawalla, S.B. and J.F. Rosenbaum, Placebo response in depression. Dialogues Clin Neurosci, 2002. 4(1): p. 105–13.

9. Rutherford, B.R. and S.P. Roose, A model of placebo response in antidepressant clinical trials. Am J Psychiatry, 2013. 170(7): p. 723–33.

10. Enck, P., et al., The placebo response in medicine: minimize, maximize or personalize? Nature Reviews Drug Discovery, 2013. 12(3): p. 191–204.

11. Fava, M., et al., The Problem of the Placebo Response in Clinical Trials for Psychiatric Disorders: Culprits, Possible Remedies, and a Novel Study Design Approach. Psychotherapy and Psychosomatics, 2003. 72(3): p. 115–127.

12. Rosenkranz, G.K., Remarks on designs enriching for placebo non-responders. Clin Trials, 2016. 13(3): p. 338–43.

13. Baskaran, A., R. Milev, and R.S. McIntyre, The neurobiology of the EEG biomarker as a predictor of treatment response in depression. Neuropharmacology, 2012. 63(4): p. 507–513.

14. Olbrich, S. and M. and Arns, EEG biomarkers in major depressive disorder: Discriminative power and prediction of treatment response. International Review of Psychiatry, 2013. 25(5): p. 604–618.

15. Thase, M.E., Using biomarkers to predict treatment response in major depressive disorder: evidence from past and present studies. Dialogues Clin Neurosci, 2014. 16(4): p. 539–44.

16. Watts, D., et al., Predicting treatment response using EEG in major depressive disorder: A machine-learning meta-analysis. Translational Psychiatry, 2022. 12(1): p. 332.

17. Widge, A.S., et al., Electroencephalographic Biomarkers for Treatment Response Prediction in Major Depressive Illness: A Meta-Analysis. American Journal of Psychiatry, 2018. 176(1): p. 44–56.

18. Jaworska, N., et al., Leveraging Machine Learning Approaches for Predicting Antidepressant Treatment Response Using Electroencephalography (EEG) and Clinical Data. Front Psychiatry, 2018. 9: p. 768.

19. Korb, A.S., et al., Rostral anterior cingulate cortex theta current density and response to antidepressants and placebo in major depression. Clin Neurophysiol, 2009. 120(7): p. 1313–9.

20. Leuchter, A.F., et al., Changes in brain function of depressed subjects during treatment with placebo. Am J Psychiatry, 2002. 159(1): p. 122–9.

21. Pizzagalli, D.A., et al., Pretreatment Rostral Anterior Cingulate Cortex Theta Activity in Relation to Symptom Improvement in Depression: A Randomized Clinical Trial. JAMA Psychiatry, 2018. 75(6): p. 547–554.

22. Bartova, L., et al., Reduced default mode network suppression during a working memory task in remitted major depression. Journal of Psychiatric Research, 2015. 64: p. 9–18.

23. Fang, Z., et al., Functional connectivity profiles in remitted depression and their relation to ruminative thinking. NeuroImage: Clinical, 2025. 45: p. 103716.

24. Kaiser, R.H., et al., Large-Scale Network Dysfunction in Major Depressive Disorder: A Meta-analysis of Resting-State Functional Connectivity. JAMA Psychiatry, 2015. 72(6): p. 603–11.

25. Yang, Z., et al., Understanding complex functional wiring patterns in major depressive disorder through brain functional connectome. Transl Psychiatry, 2021. 11(1): p. 526.

26. Wang, L., et al., A systematic review of resting-state functional-MRI studies in major depression. J Affect Disord, 2012. 142(1-3): p. 6–12.

27. Chin Fatt, C.R., et al., Data driven clusters derived from resting state functional connectivity: Findings from the EMBARC study. J Psychiatr Res, 2023. 158: p. 150–156.

28. Jiao, Y., et al., Deep graph learning of multimodal brain networks defines treatment-predictive signatures in major depression. Mol Psychiatry, 2025.

29. Chin Fatt, C.R., et al., Differential response to SSRI versus Placebo and distinct neural signatures among data-driven subgroups of patients with major depressive disorder. J Affect Disord, 2021. 282: p. 602–610.

30. Drysdale, A.T., et al., Resting-state connectivity biomarkers define neurophysiological subtypes of depression. Nat Med, 2017. 23(1): p. 28–38.

31. Schmaal, L., et al., Predicting the Naturalistic Course of Major Depressive Disorder Using Clinical and Multimodal Neuroimaging Information: A Multivariate Pattern Recognition Study. Biol Psychiatry, 2015. 78(4): p. 278–86.

32. Trivedi, M.H., et al., Establishing moderators and biosignatures of antidepressant response in clinical care (EMBARC): Rationale and design. J Psychiatr Res, 2016. 78: p. 11–23.

33. Li Q, D.M., Breier A, Potter W, et al., AN EEG-BASED, MACHINE LEARNING BIOMARKER TO IDENTIFY RESPONSIVE VS. NON-RESPONSIVE SUBJECTS IN AN MDD CLINICAL TRIAL: INITIAL VALIDATION DATA FROM THE EMBARC STUDY DATABASE, in International Society for CNS Clinical Trials and Methodology (ISCTM) 2024: Washington, DC.

34. Etkin, A. and D.H. Mathalon, Bringing Imaging Biomarkers Into Clinical Reality in Psychiatry. JAMA Psychiatry, 2024. 81(11): p. 1142–1147.

35. Rette, D., et al., Neural Predictors of the Antidepressant Placebo Response. Pharmaceuticals (Basel), 2019. 12(4).

36. Abi-Dargham, A., et al., Candidate biomarkers in psychiatric disorders: state of the field. World Psychiatry, 2023. 22(2): p. 236–262.

37. García-Gutiérrez, M.S., et al., Biomarkers in Psychiatry: Concept, Definition, Types and Relevance to the Clinical Reality. Front Psychiatry, 2020. 11: p. 432.

38. Barreiros, A.R., et al., Intra- and Inter-Network connectivity of the default mode network differentiates Treatment-Resistant depression from Treatment-Sensitive depression. Neuroimage Clin, 2024. 43: p. 103656.

39. Whitton, A.E., et al., Pretreatment Rostral Anterior Cingulate Cortex Connectivity With Salience Network Predicts Depression Recovery: Findings From the EMBARC Randomized Clinical Trial. Biol Psychiatry, 2019. 85(10): p. 872–880.

40. Rolle, C.E., et al., Cortical Connectivity Moderators of Antidepressant vs Placebo Treatment Response in Major Depressive Disorder: Secondary Analysis of a Randomized Clinical Trial. JAMA Psychiatry, 2020. 77(4): p. 397–408.

41. He, B., et al., Electrophysiological Source Imaging: A Noninvasive Window to Brain Dynamics. Annu Rev Biomed Eng, 2018. 20: p. 171–196.

42. Warbrick, T., Simultaneous EEG-fMRI: What Have We Learned and What Does the Future Hold? Sensors (Basel), 2022. 22(6).

43. Klooster, D., et al., Evaluating Robustness of Brain Stimulation Biomarkers for Depression: A Systematic Review of Magnetic Resonance Imaging and Electroencephalography Studies. Biological Psychiatry, 2024. 95(6): p. 553–563.

44. Lopez, K.L., et al., Stability, change, and reliable individual differences in electroencephalography measures: A lifespan perspective on progress and opportunities. NeuroImage, 2023. 275: p. 120116.

45. Zhang, Y., et al., Identification of psychiatric disorder subtypes from functional connectivity patterns in resting-state electroencephalography. Nature Biomedical Engineering, 2021. 5(4): p. 309–323.

46. Schwartzmann, B., et al., Developing an Electroencephalography-Based Model for Predicting Response to Antidepressant Medication. JAMA Netw Open, 2023. 6(9): p. e2336094.

47. Tong, X., et al., Individual deviations from normative electroencephalographic connectivity predict antidepressant response. J Affect Disord, 2024. 351: p. 220–230.

